# Adult mortality before and during the COVID-19 pandemic in nine communities of Yemen: a key informant study

**DOI:** 10.1101/2022.06.20.22276574

**Authors:** Mervat Alhaffar, Huda BaSaleem, Fouad Othman, Khaled Alsakkaf, Sena Mohammed Mohsen Alkhteeb, Hussein Kolaise, Abdullah K. Babattah, Yaseen Abdulmalik Mahyoub Salem, Hannah Brindle, Najwa Yahya, Pasquale Pepe, Francesco Checchi

**Affiliations:** Department of Infectious Disease Epidemiology, Faculty of Epidemiology and Population Health, London School of Hygiene and Tropical Medicine, London, UK; Department of Community Medicine and Public Health, Faculty of Medicine and Health Science, University of Aden, Aden, Yemen; Faculty of Medicine and Health Science, Ta’iz University, Ta’iz, Yemen; Epidemiology Surveillance Office, Ta’iz Governorate Health Office, Ta’iz, Yemen; Department of Internal Medicine, Faculty of Medicine and Health Science, University of Aden, Aden, Yemen; Primary Health Care Program, Health Sector, HUMAN ACCESS for Partnership and Development, Aden, Yemen; Yemen Public Health Network, Aden, Yemen

**Keywords:** Yemen, humanitarian, armed conflict, crisis, mortality, SARS-CoV-2, COVID-19, key informant, capture recapture, multiple systems estimation

## Abstract

**Introduction:** Widespread armed conflict has affected Yemen since 2014. To date, the mortality toll of seven years of crisis, and any excess due to the COVID-19 pandemic, are not well quantified. We attempted to estimate population mortality during the pre-pandemic and pandemic periods in nine purposively selected urban and rural communities of southern and central Yemen (Aden and Ta’iz governorates), totalling > 100,000 people.

**Methods:** Within each study site, we collected lists of decedents between January 2014-March 2021 by interviewing different categories of key community informants, including community leaders, imams, healthcare workers, senior citizens and others. After linking records across lists based on key variables, we applied two-, three- or four-list capture-recapture analysis to estimate total death tolls. We also computed death rates by combining these estimates with population denominators, themselves subject to estimation.

**Results:** After interviewing 138 disproportionately (74.6%) male informants, we identified 2445 unique decedents. While informants recalled deaths throughout the study period, reported deaths among children were sparse: we thus restricted analysis to persons aged ≥15 years old. We noted a peak in reported deaths during May-July 2020, plausibly coinciding with the first COVID-19 wave. Death rate estimates featured uninformatively large confidence intervals, but appeared elevated compared to the non-crisis baseline, particularly in two sites where a large proportion of deaths were attributed to war injuries. There was no clear-cut evidence of excess mortality during the pandemic period.

**Conclusions:** We found some evidence of a peak in mortality during the early phase of the pandemic, but death rate estimates were otherwise too imprecise to enable strong inference on trends. Estimates suggested substantial mortality elevations from baseline during the crisis period, but are subject to serious potential biases. The study highlighted challenges of data collection in this insecure, politically contested environment.

## Background

Yemen has been affected by political unrest throughout recent history [1–3]. The current armed conflict started in 2014. The country is split into several areas of control with contested borders and multiple local and international regional actors holding different political and ideological stances [3–6]. The protracted armed conflict resulted in targeting civilians, healthcare facilities and personnel, water and sanitation infrastructure, schools, roads and bridges thereby disrupting services and exacerbating poor living conditions experienced before the war [5, 7–10]. In addition to frequent civilian casualties, the protracted conflict has resulted in widespread forced displacement, food insecurity, severe malnutrition and food insecurity bordering on potential famine [11], and a heavily disrupted and fragmented health system consisting of two ministries of health (controlled by two different authorities in the north and the south), one based in Sana’a and the other in Aden [8, 12].

The first laboratory-confirmed case of COVID-19 in Yemen was announced in April 2020 [13]. Efforts to track the pandemic’s dynamics and share information relating to its impact on morbidity and mortality have differed between areas of political control. Houthi authorities in particular have tended not to openly report cases and deaths in their areas of control [14–16].

In conflict settings, information on health status can help humanitarian actors better allocate scarce resources and record the progress of response activities [17, 18]. Population mortality is a key indicator of health status as it captures the contributions of various health risk factors [18, 19]. Additionally, documenting mortality during conflicts may support advocacy for peacebuilding and post-conflict reconciliation [20].

To contribute evidence on COVID-19 attributable mortality in Yemen, we aimed to estimate the excess death toll and death rate during the pandemic period, as well as the pre-pandemic levels, in nine selected sites in the south and central of the country by collecting lists of decedents from key community informants, and applying capture-recapture statistical analysis to these. A secondary aim was to explore the feasibility and limitations of conducting such research in this context.

## Methods

### Scoping phase

Prior to data collection, we conducted a scoping phase consisting of informal in-depth interviews and group discussions in Arabic with more than seventy people who identified as Yemenis in the United Kingdom (UK), Yemen, and other countries. The participants were identified through a snowball approach of sequential referrals while ensuring a difference in their backgrounds including profession and place of origin in Yemen. The interviews took place in person in London and Sheffield (after obtaining verbal consent), and online with note-taking used to record the information collected. Interviews included questions on (i) the categories of people most likely to know about deaths and births in the community, (ii) the ways in which people report/know about deaths in the community, (iii) whether health facilities or burial sites keep any written lists on deceased people and (iv) security concerns that might impact participation or bias people’s answers in different areas in Yemen. This scoping phase helped us to form an independent advisory group of country-based Yemeni researchers and public health actors to provide guidance on the project, and establish collaborations between the London School of Hygiene and Tropical Medicine, the University of Aden and Ta’iz University.

### Study population and period

While we originally wished to select a nationally representative sample, study permission was not received from authorities in Sana’a (the Supreme Council for the Management and Coordination of Humanitarian Affairs and International Cooperation SCMCHA)[21], and even in southern Yemen, the geographical scope of data collection was constrained by travel and security challenges. We thus opted to purposively select nine accessible sites including four in Aden governorate and five in Ta’iz governorate which were identified by Yemen-based authors and represented different levels and typologies of exposures to the crisis (Table 1). We further selected sites that were clearly delineated (e.g. city neighbourhoods, groups of villages) and of an approximate population size (5000-20,000) consistent with feasible data collection within the study’s resources and security constraints. All the sites were under the control of the Internationally Recognised Government (IRG), site names are omitted here to maintain confidentiality of the key informants. Data collection started on 18^th^ January and ended on 31^st^ March 2021. We inquired about deaths among people residing within the site from January 2014 to the start of data collection.

**Table 1.** Characteristics of the study sites and informant lists retained for capture-recapture analysis. A and T refer to sites in Aden and Ta’iz governorates, respectively.

| Site | Rural/<br>Urban | Characteristics<br>(estimated population, March 2021) | Informant lists<br>(main analysis) | Informant lists<br>(alternative analysis) |
| --- | --- | --- | --- | --- |
| A1 | Urban | Directly and heavily affected by the armed conflict; under Houthi control for around four months. Residents fled the area due to the lack of water, electricity and fuel, and shortage of food and health services.<br>(population size uncertain) | 1. Community leaders<br>2. Health workers<br>3. Imams<br>4. Burial preparers | 1. Community leaders<br>2. Health workers<br>3. Imams + Burial preparers |
| A2 | Urban | Directly and heavily affected by the armed conflict; under Houthi control for around four months. Residents fled the area due to the lack of water, electricity and fuel, and shortage of food and health services.<br>(population 11,000, ≈9% of subdistrict) | 1. Community leaders<br>2. Senior citizens<br>3. Imams<br>4. Burial preparers | 1. Community leaders + senior citizens<br>2. Imams + Burial preparers |
| A3 | Urban | Moderately affected by the armed conflict; destination of many internally displaced populations (IDPs).<br>(population 2600, ≈2% of subdistrict) | 1. Community leaders<br>2. Imams<br>3. Burial preparers | 1. Community leaders<br>2. Imams + Burial preparers |
| A4 | Urban | Moderately affected by the armed conflict; destination of many IDPs.<br>(population 9400, ≈6% of subdistrict) | 1. Community leaders + spouses<br>2. Senior citizens<br>3. Burial preparers | 1. Community leaders + spouses + senior citizens<br>2. Burial preparers |
| T1 | Urban | On the contested borders between the two areas of political control.<br>(population 2700, ≈3% of subdistrict) | 1. Community leaders + spouses + senior citizens + imams<br>2. Health workers | 1. Community leaders + spouses + senior citizens<br>2. Health workers<br>3. Imams + Burial preparers |
| T2 | Rural area | Ongoing direct conflict since the beginning of the crisis; divided between two areas of control.<br>(population 14,700 100% of subdistrict) | 1. Community leaders + senior citizens + imams<br>2. Health workers<br>3. Teachers<br>4. Burial preparers | 1. Community leaders + senior citizens + teachers<br>2. Health workers<br>3. Imams + burial preparers |
| T3 | Semi-urban | No active conflict, but a destination for many IDPs.<br>(population size uncertain) | 1. Community leaders + spouses + senior citizens + imams<br>2. Health workers | 1. Community leaders + spouses + senior citizens<br>2. Health workers<br>3. Imams |
| T4 | Urban | Past direct conflict, was under bombardment and siege.<br>(population 8600, ≈15% of subdistrict) | 1. Community leaders + senior citizens<br>2. Health workers | 1. Community leaders + senior citizens<br>2. Health workers |
| T5 | Rural area | Ongoing direct conflict since the beginning of the crisis, divided between two areas of control.<br>(population 21,700, 100% of subdistrict) | 1. Community leaders<br>2. Senior citizens | 1. Community leaders + senior citizens<br>(no analysis possible: only one list) |

Several key informant categories were identified: community leaders and their spouses; healthcare providers; imams; ritual burial preparers; senior citizens; and teachers. None were known to keep written death records, except for health facility managers. Moreover, in places where the conflict was intense, records from local hospitals were destroyed or were missing data from specific years or were not even collected. Depending on the site’s characteristics e.g. urban versus rural or size, some or all of the above informant categories were identifiable and thus included in the study.

### Participant recruitment

The in-country research team consisted of three male medical doctors and one female midwife who were residents and familiar with the study communities. They initially drew on their personal and professional connections to identify key informants and build trust before conducting the interviews. Participants provided verbal, researcher-witnessed informed consent. Those who declined to participate were mostly able to recommend others to be interviewed. All key informants provided consent in-person, mostly after receiving recommendations from respected peers, e.g. another community leader or a hospital manager, who either wrote letters of introduction or accompanied researchers on their first visits to potential participants. In Aden, preliminary phone conversations helped to establish rapport with key informants and facilitate organizing appointments for introducing the study while in Ta’iz in-person visits were necessary. In Aden 13/69 people contacted (two women and 11 men) declined to participate in the study, while in Ta’iz 7/89 (all men) declined. Stated reasons for non-participation included concerns that information would be shared with opposing forces; negative previous experiences with researchers; insufficient time; and anticipated difficulty to recall deaths.

### Data collection

A standardised questionnaire was developed and built using the Open Data Kit (ODK) [22] platform, with in-built validation rules to collect information on (i) key informants (age, gender, typology, site and residency) and (ii) names, age, gender, year of death, cause of death, place of death and origin of decedents within the sites since 2014. Online two-day training sessions for researchers took place in Arabic consisting of study concepts, procedures for data collection and management, and role-play. Data were entered into the ODK Application on Android devices and automatically uploaded to a secure server hosted at the LSHTM. Participants were also offered the option to recall and write down decedent information in their own time on a printed form, if needed enlisting help from their spouses; researchers collected the form at a later point and entered the data into ODK.

Investigators in Aden, Ta’iz and London monitored data quality, addressed issues and replied to questions in real-time. After conducting 56 interviews, we paused data collection for a week to share experiences. Given that the preliminary analysis showed a disproportionate reporting of adult male deaths, we agreed on procedures to enhance the detection of child and women decedents during subsequent interviews. This included a heavier reliance on spouses for recall and listing female names as ‘mother/wife/daughter of’, which was felt to be more protective of their confidentiality.

### Analysis

#### Record linkage

Data were manually cleaned, and duplicates removed from each key informant’s list. Linkage criteria (Table S1, Supplementary File) were developed to establish, for each site, which lists a given decedent was included in. Individuals’ names in Yemen usually consist of first name, father’s name, grandfather’s name and lastly tribe name or area/ district of origin. Records with missing first name, gender, age, and/or year of death were excluded from analysis. The year and age at death were averaged if informants reported discordant values.

#### Capture-recapture analysis

Capture-recapture analysis examines the overlap among lists *L* to estimate the number of individuals (in this case decedents) who have not been captured by any list. This estimate, summed to the number of individuals appearing on at least one list, provides the total. In this study, lists consisted for each site of the records collected from specific categories of informants. After record linkage, site data consisted of two, three or four lists (see below and Table 1).

In a two-list scenario, each decedent *x* ∈ {1,2,3 … *N*} has status *x*_10_ if named within *L*_1_ only, *x*_01_ if named within *L*_2_ only, *x*_11_ if named by both lists and *x*_00_ if not captured by either list. The resulting contingency table consists of four cells, *n*_10_, *n*_01_, *n*_11_ and *n*_00_, the last of which is unknown. We used the simple Chapman estimator to estimate the total number of deaths as 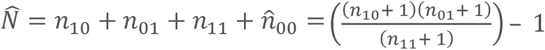; we computed a confidence interval (CI) as 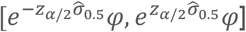, where 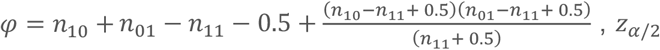 is the normal distribution quantile for a given significance level of interest (1.96 for *α* = 0.05 or 95%CI) and 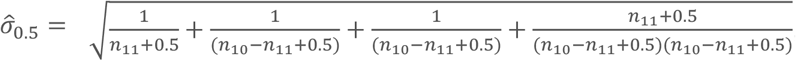, as per Sadinle [23].

In a three-list scenario, the overlap among lists *L* may be represented by eight alternative candidate log-linear Poisson models, each of which features terms for the probability of appearing on any given list, as well as two-way interaction terms representing potential dependencies among lists: these models range from one with no interaction terms to a model featuring all the two-way interactions *L*_1_ × *L*_2_, *L*_2_ × *L*_3_ and *L*_1_ × *L*_3_. We wished to also include in the models an exposure (period before and during the COVID-19 pandemic in Yemen) and potential confounding variables (age, gender). To allow for continuous covariates, we used Rossi et al.’s [24] parametrisation of log-linear models, whereby the dataset is expanded to feature, for each individual, all potential list statuses (*x*_000,_ *x*_100,_ *x*_101_, *x*_001_, *x*_110_, *x*_101_, *x*_011_, *x*_111_); an outcome of 1 for the actual status, missing for status *x*_000_ and 0 otherwise; and any covariate values. The model, once fit, is used to predict 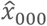, interpretable as each individual’s contribution to 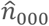, the estimate of uncaptured deaths (i.e. 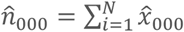); this quantity may of course be stratified by exposure stratum. This estimation framework can easily be extended to the four-list scenario, which however entails a larger set of models, featuring both three-list and/or (hierarchically non-redundant) two-list interactions. While conventional capture-recapture analysis selects the best-fitting among candidate models, we adapted Rossi et al.’s suggested approach for averaging multiple models [25]. First, we screened out models that did not fit (e.g. due to sparse overlap among lists), yielded an implausible 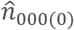 (defined as ≥ 10 times the number of listed deaths) or featured a likelihood-ratio test p-value ≥ 0.60 when compared to the saturated model (indicating potential overfitting). At this stage we also assessed whether to retain any potential confounder covariates, based on likelihood-ratio tests compared to the no-confounder model and inspection of estimates with and without the confounder. For each shortlisted model *i* ∈ {1,2,3 … *K*}, we then computed a weight (equivalent to a Bayesian posterior probability) between 0 and 1 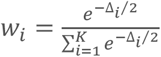, where Δ_*i*_= *AIC*_*i*_ − *AIC*_min_, i.e. the difference between the model’s Akaike Information Criterion (AIC) and the lowest AIC among all shortlisted models (the AIC is a goodness-of-fit indicator that rewards predictive accuracy and parsimony, i.e. the fewest possible model terms). Finally, we computed a weighted average estimate of 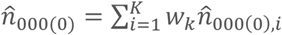. We present results overall and by period.

#### Alternative list groupings

Capture-recapture analysis is unfeasible when the overlap among the lists is very poor, causing models not to fit (see above). To avoid these problems, for our main analysis we grouped lists together that plausibly reflected similar sources of community information: community leaders’ and their spouses’ lists were combined in sites A4, T1 and T3; community leaders’ and senior citizens’ lists were combined in sites T1, T2, T3 and T4; community leaders’ and imams’ lists were combined in sites T1, T2 and T3. This yielded between two and four grouped lists per site (Table 1).

As an alternative analysis, we (i) combined imams’ and burial preparers’ lists across all sites to form one list and (ii) added records obtained from teachers and senior citizens to the community leaders’ lists.

### Population estimates and death rates

We previously estimated the all-age population of Yemen between 2014 and 2021, by month and sub-district (administrative level 3), using a combination of pre-crisis census, geospatial projections by WorldPop (available at 100 m^2^ resolution, and resulting from extensively validated predictive statistical models [26]) and displacement flow data. Details are provided in Checchi et al. [27]. However, only two sites (T2, T5) consisted of entire subdistricts. In remaining sites, researchers attempted to collect GPS coordinates of all corners of the site (e.g. road boundaries) using their phones. We overlaid these polygons, and the boundaries of the surrounding subdistricts, onto WorldPop projections for 2017 (the mid-point of the data collection period) to compute the approximate proportion of the subdistrict’s population that fell within the site. We then multiplied our subdistrict estimates by this proportion to estimate the site’s population. Site boundaries, and thus populations, were unresolved for two sites (A1, T3). We used population estimates to compute average death rates by period (pre-pandemic and pandemic). We specifically present crude death rates among people aged ≥15yo (CDR15+), per all-age population: this may be thought of as an age-specific fraction of the all-age CDR.

## Results

### Decedent records

Between 14 January and 31 March 2021, 138 interviews were conducted (56 in Aden, 82 in Ta’iz: Figure 1), yielding 3022 decedent records. Across all sites, only 35/138 (25.4%) interviews were conducted with female informants (midwives, health workers, wives of community leaders and burial preparers).

**Figure 1.**
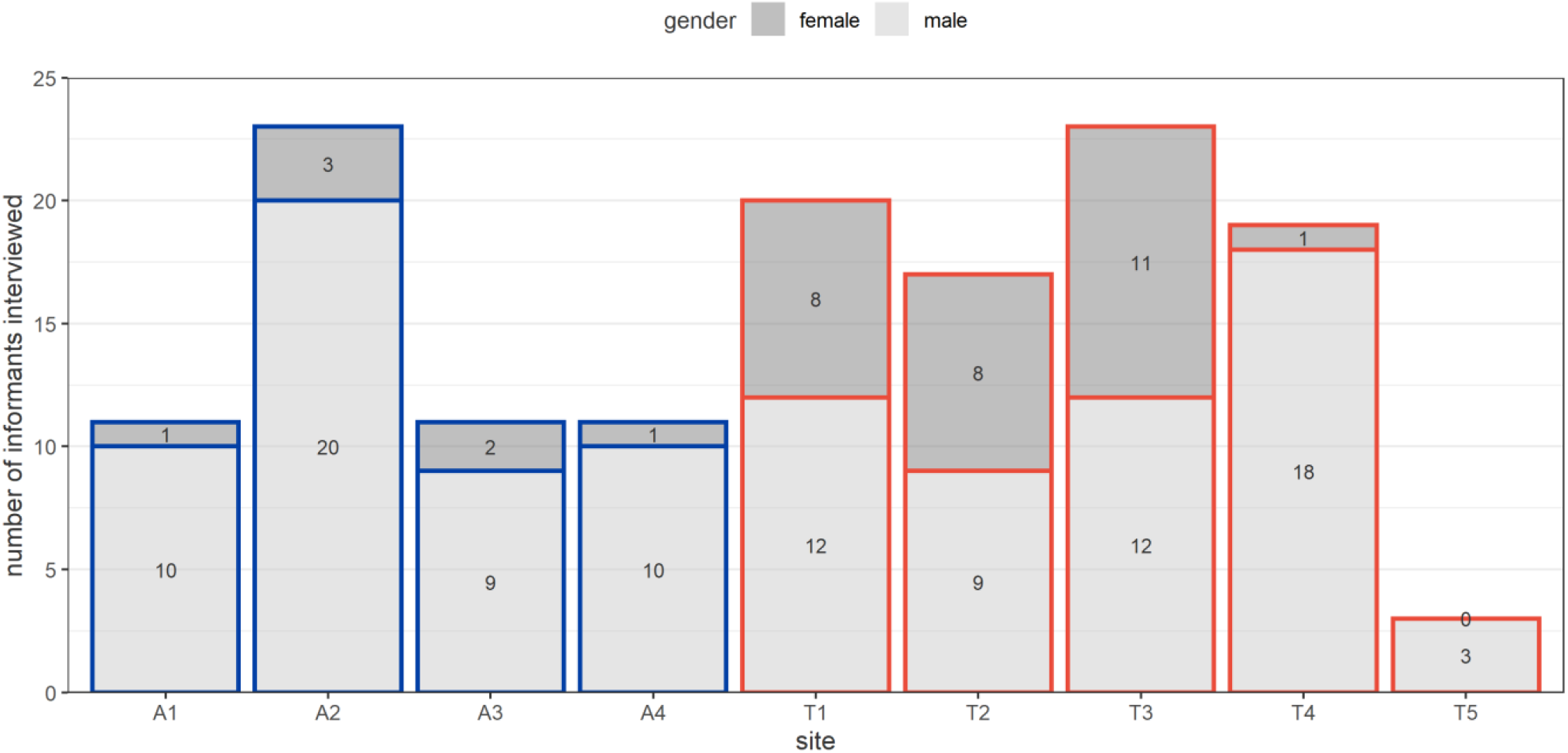
Number of key informant interviews done, by site and gender of the informant.

Apart from 2014, when the number of recalled deaths was 138, the annual number of death events recalled by informants did not appear to decrease as time became more remote (range = 269 to 372) (Figure 2). The largest number of deaths were reported for 2020 (N = 574). The highest number of deaths was reported by participants from site T1 (N = 718) and the lowest from T5 (N = 81).

**Figure 2.**
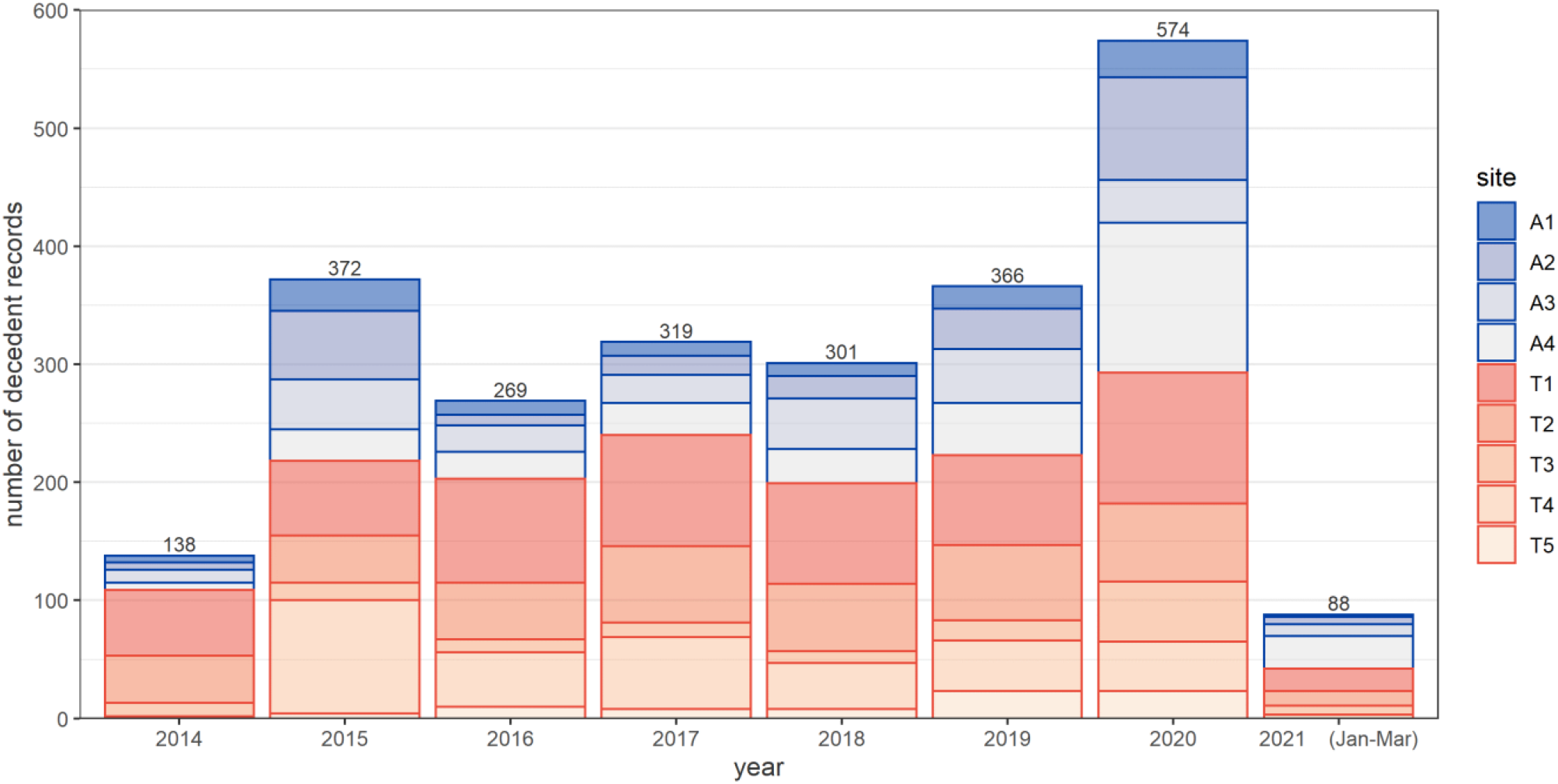
Number of decedent records reported by key informants, by year and site.

### Characteristics of reported deaths

After record linkage (including definite, probable and possible matches) and excluding 21 non-analysable records due to one or more missing variables, 2445 unique decedents were identified across all sites. All sites except for T3 were notable for the sparsity of data on children (only 189/2445 or 7.7% reported decedents were aged <15yo), and for a disproportion of males (1851/2445, 75.7%; Table 2). Site T4 featured an exceptionally large share of deaths in the 15-44yo stratum while in the remaining sites the highest numbers of deaths were reported among people aged ≥ 45yo. Site-specific detail by list is provided in the Supplementary File.

**Table 2.** Age and gender distribution of reported deaths, by site.

| site: | A1 | A2 | A3 | A4 | T1 | T2 | T3 | T4 | T5 |
| --- | --- | --- | --- | --- | --- | --- | --- | --- | --- |
| **Age** |  |  |  |  |  |  |  |  |  |
| Mean | 48.5y | 57.3y | 54.4y | 58.1y | 43.6y | 49.7y | 38.1y | 33.4y | 53.9y |
| 0y | 1<br>(0.8%) | 0<br>(0.0%) | 2<br>(0.9%) | 2<br>(0.6%) | 26<br>(4.3%) | 9<br>(2.3%) | 27<br>(20.0%) | 4<br>(1.2%) | 0<br>(0.0%) |
| 1y to 4y | 3<br>(2.5%) | 2<br>(0.9%) | 1<br>(0.4%) | 0<br>(0.0%) | 6<br>(1%) | 11<br>(2.8%) | 7<br>(5.2%) | 3<br>(0.9%) | 0<br>(0.0%) |
| 5y to 14y | 6<br>(5.0%) | 3<br>(1.3%) | 1<br>(0.4%) | 6<br>(1.9%) | 29<br>(4.8%) | 22<br>(5.6%) | 6<br>(4.4%) | 10<br>(3.0%) | 2<br>(2.6%) |
| 15y to 44y | 37<br>(30.8%) | 44<br>(18.7%) | 64<br>(27.4%) | 65<br>(20.9%) | 252<br>(41.3%) | 111<br>(28.3%) | 35<br>(25.9%) | 244<br>(73.5%) | 19<br>(25.0%) |
| $\geq 45$y | 73<br>(60.8%) | 186<br>(79.1%) | 166<br>(70.9%) | 238<br>(76.5%) | 297<br>(48.7%) | 239<br>(61%) | 60<br>(44.4%) | 71<br>(21.4%) | 55<br>(72.4%) |
| **Gender** |  |  |  |  |  |  |  |  |  |
| Female | 36<br>(30.0%) | 70<br>(29.8%) | 53<br>(22.6%) | 86<br>(27.7%) | 115<br>(18.9%) | 132<br>(33.8%) | 45<br>(33.3%) | 31<br>(9.3%) | 26<br>(34.2%) |
| Male | 84<br>(70.0%) | 165<br>(70.5%) | 181<br>(77.7%) | 225<br>(72.8%) | 495<br>(81.1%) | 260<br>(66.3%) | 90<br>(66.7%) | 301<br>(90.7%) | 50<br>(65.8%) |
| Totals | 120 | 235 | 234 | 311 | 610 | 392 | 135 | 332 | 76 |

For 2020 and 2021 only, we collected months of death: when aggregating all sites, a notable peak in monthly deaths is evident in May-July 2020, possibly corresponding to the first COVID-19 wave (Figure 3), with a majority of recalled deaths among people aged ≥ 45yo.

**Figure 3.**
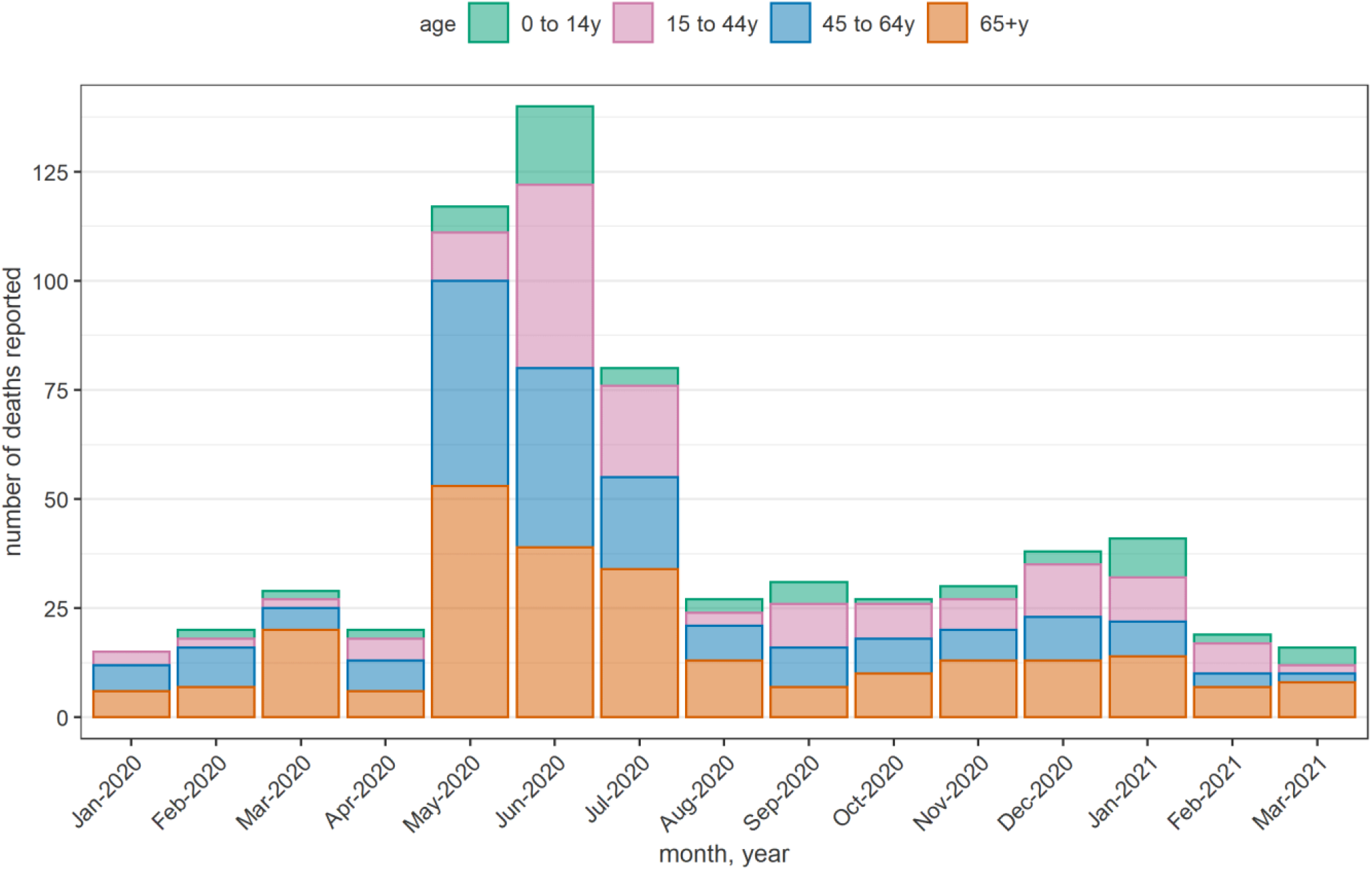
Number of unique decedents per month reported by informants across all sites, January 2020 to March 2021.

Across all sites and particularly in A1, T1 and T4, a substantial proportion of deaths was attributed to intentional or war-related injuries (Table 3).

**Table 3.** Causes of death as reported by informants over the entire analysis period, by site.

| Site | Intentional/war injury | Unintentional injury | Other causes | Total |
| --- | --- | --- | --- | --- |
| A1 | 44 (36.7%) | 15 (12.5%) | 61 (50.8%) | 120 |
| A2 | 38 (16.2%) | 3 (1.3%) | 194 (82.6%) | 235 |
| A3 | 46 (19.7%) | 15 (6.4%) | 173 (73.9%) | 234 |
| A4 | 50 (16.1%) | 11 (3.5%) | 250 (80.4%) | 311 |
| T1 | 244 (40.0%) | 31 (5.1%) | 335 (54.9%) | 610 |
| T2 | 61 (15.6%) | 23 (5.9%) | 308 (78.6%) | 392 |
| T3 | 11 (8.1%) | 11 (8.1%) | 113 (83.7%) | 135 |
| T4 | 168 (50.6%) | 106 (31.9%) | 58 (17.5%) | 332 |
| T5 | 12 (15.8%) | 0 (0.0%) | 64 (84.2%) | 76 |

### Estimates of total adult mortality

Across all sites, estimates of total deaths overall and by period before and after the first COVID-19 case was announced in Yemen featured wide 95% confidence intervals (95%CI; Table 4). When considering point estimates, the mean monthly mortality rose appreciably during March 2020 to March 2021 compared to January 2014 to February 2020 in sites A1, A2, A4, T3 and T5, remained nearly constant in A3, T1 and T2, and declined in T4. The alternative analysis yielded an approximately similar pattern (Table 5).

**Table 4.**
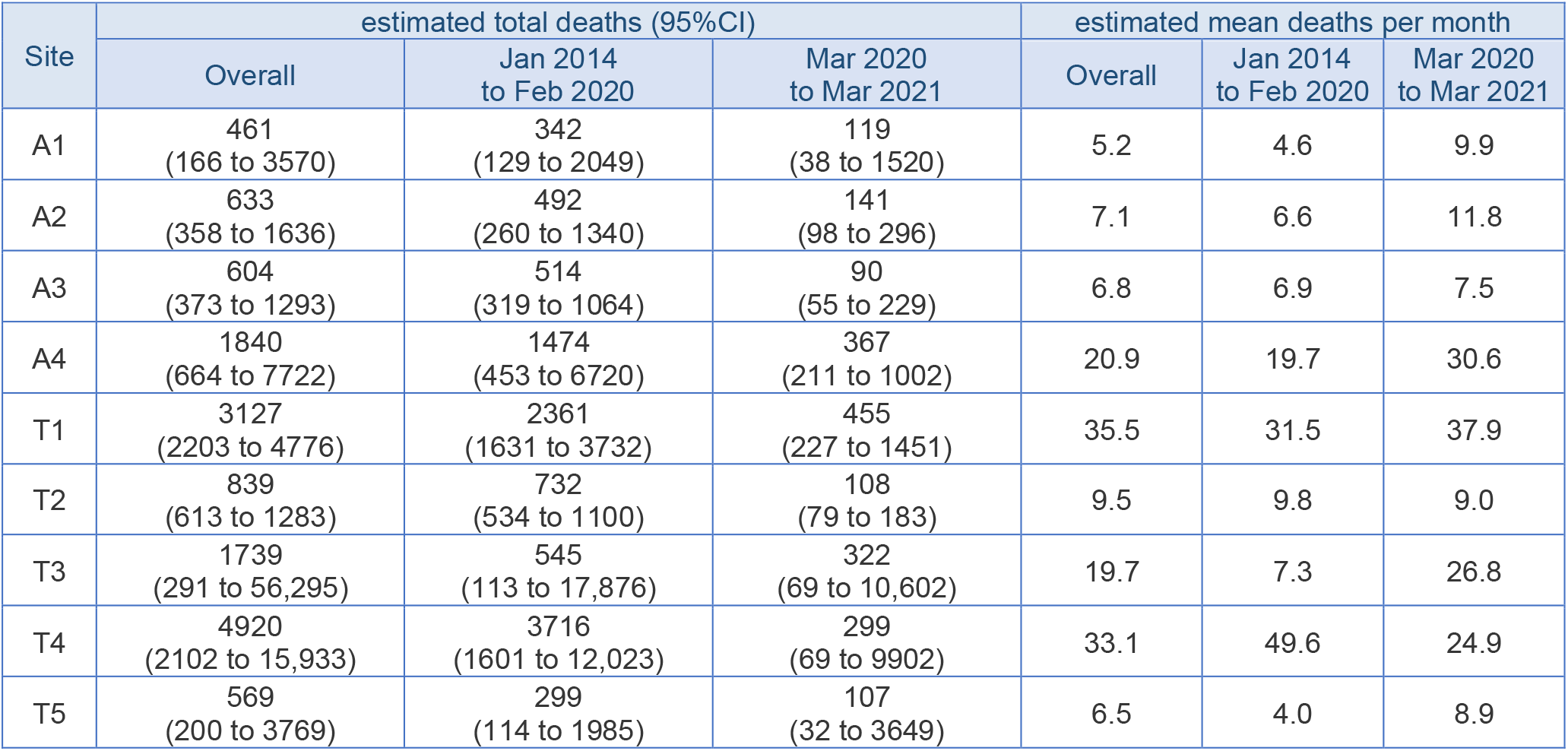
Estimates of total deaths and deaths per month, overall and by period (main analysis).

**Table 5.** Estimates of total deaths and deaths per month, overall and by period (alternative analysis).

| Site | estimated total deaths (95%CI) |  |  | estimated mean deaths per month |  |  |
| --- | --- | --- | --- | --- | --- | --- |
|  | Overall | Jan 2014 to Feb 2020 | Mar 2020 to Mar 2021 | Overall | Jan 2014 to Feb 2020 | Mar 2020 to Mar 2021 |
| A1 | 207<br>(35 to 1703) | 155<br>(28 to 1158) | 52<br>(7 to 544) | 2.3 | 2.1 | 4.3 |
| A2 | 441<br>(368 to 567) | 308<br>(243 to 437) | 132<br>(106 to 196) | 5.0 | 4.1 | 11.0 |
| A3 | 534<br>(424 to 733) | 448<br>(346 to 645) | 79<br>(56 to 169) | 6.1 | 6.0 | 6.6 |
| A4 | 845<br>(569 to 1587) | 818<br>(322 to 5430) | 296<br>(211 to 568) | 9.6 | 10.9 | 24.7 |
| T1 | No results (all models yielded implausible estimates) |  |  |  |  |  |
| T2 | 915<br>(587 to 2060) | 802<br>(509 to 1815) | 113<br>(78 to 244) | 10.4 | 10.7 | 9.4 |
| T3 | No results (all models yielded implausible estimates) |  |  |  |  |  |
| T4 | 4126<br>(2074 to 10,274) | 4719<br>(2019 to 15,279) | 119<br>(64 to 485) | 46.8 | 62.9 | 9.9 |
| T5 | No results (only one list) |  |  |  |  |  |

Available CDR15+ estimates varied considerably across sites (Table 6). There was no consistent pattern of increase during the pandemic period, though 95%CIs are uninformatively wide. Sites T1 and T4, where the highest CDR15+ was observed, were also those with the highest percentage of intentional/war injury deaths (Table 3) and deaths among young adults (Table 2).

**Table 6.** Estimated crude death rates among people aged ≥15yo, overall and by period (main analysis).

| Site | deaths ≥15yo per 1000 all-age population per year (95%CI) |  |  | deaths ≥15yo per 10,000 all-age population per day (95%CI) |  |  |
| --- | --- | --- | --- | --- | --- | --- |
|  | Overall | Jan 2014 to Feb 2020 | Mar 2020 to Mar 2021 | Overall | Jan 2014 to Feb 2020 | Mar 2020 to Mar 2021 |
| A1 | [no population estimates] |  |  |  |  |  |
| A2 | 9.1<br>(5.2 to 23.5) | 8.5<br>(4.5 to 23.2) | 11.9<br>(8.3 to 25.0) | 0.2<br>(0.1 to 0.6) | 0.2<br>(0.1 to 0.6) | 0.3<br>(0.2 to 0.7) |
| A3 | 38.3<br>(23.7 to 82) | 39.5<br>(24.5 to 81.7) | 32.8<br>(20.0 to 83.4) | 1.0<br>(0.6 to 2.2) | 1.1<br>(0.7 to 2.2) | 0.9<br>(0.5 to 2.3) |
| A4 | 28.2<br>(10.2 to 118.3) | 27.3<br>(8.4 to 124.3) | 32.7<br>(18.8 to 89.4) | 0.8<br>(0.3 to 3.2) | 0.7<br>(0.2 to 3.4) | 0.9<br>(0.5 to 2.4) |
| T1 | 132.9<br>(93.6 to 203) | 138<br>(95.4 to 218.2) | 70.8<br>(35.3 to 225.9) | 3.6<br>(2.6 to 5.6) | 3.8<br>(2.6 to 6.0) | 1.9<br>(1.0 to 6.2) |
| T2 | 7.5<br>(5.5 to 11.5) | 8.6<br>(6.3 to 12.9) | 4.0<br>(2.9 to 6.8) | 0.2<br>(0.2 to 0.3) | 0.2<br>(0.2 to 0.4) | 0.1<br>(0.1 to 0.2) |
| T3 | [no population estimates] |  |  |  |  |  |
| T4 | 56.0<br>(23.9 to 181.4) | 59.8<br>(25.7 to 193.3) | 11.7<br>(2.7 to 385.9) | 1.5<br>(0.7 to 5.0) | 1.6<br>(0.7 to 5.3) | 0.3<br>(0.1 to 10.6) |
| T5 | 3.8<br>(1.3 to 24.9) | 2.4<br>(0.9 to 15.9) | 4.0<br>(1.2 to 137.6) | 0.1<br>(0.0 to 0.7) | 0.1<br>(0.0 to 0.4) | 0.1<br>(0.0 to 3.8) |

## Discussion

### Main findings

Covering a population of over 100,000 and a period of seven years, our analysis suggests that nine purposively selected communities in Yemen, featuring varying exposure to the crisis, experienced highly variable death rates among people aged ≥15yo over periods before and during the COVID-19 pandemic. Due to sparse reporting of data on children, we restricted analysis to older age groups; accordingly, our CDR15+ indicators are not readily comparable with other crisis-affected settings. Based on United Nations demographic projections not accounting for crisis conditions, Yemen would have expected about 115,000 annual deaths ≥15yo during 2016-2020 within a mid-period population of 27,835,000, yielding a ‘baseline’ CDR15+ of 4.1 per 1000 person-years (0.1 per 10,000 person-days) [28]. In six of the seven study sites with available population denominators, the 95%CIs of our CDR15+ estimates exceed this baseline during all study periods, suggesting a broad pattern of considerably elevated mortality during the crisis period, with alarmingly high levels in two sites (T1, T4) where the age distribution (15-44 yo) and cause of reported deaths suggests elevated mortality directly due to armed conflict. We did not observe an obvious increase in mortality (CDR15+) during the COVID-19 transmission period, but the inaccuracy of period estimates hampers this comparison; it is possible that in some sites, extreme elevations in mortality during the early phase of the war, followed by a less acute situation, would have counterbalanced any increase due to COVID-19. The distribution of monthly reported deaths during 2020 does suggest the study sites experienced an acute epidemic peak in May-July temporally consistent with the first wave of the pandemic. A second wave was reported in Yemen shortly after this study’s data collection period.

Based on a PubMed search with keywords “mortality” and “Yemen”, we identified only two other reports of population mortality in the country since the crisis began in 2014. In Aden, satellite imagery of cemeteries and civil registration data suggested a very similar peak in mortality as in this study during May-July 2020 [13]. Ogbu et al. analysed 56 surveys done by humanitarian actors during 2015-2019 and classified Yemen’s governorates into high- and low-child mortality groups [29]; these surveys have been criticised for under-estimation [5].

The sites we investigated are unlikely to be representative of Aden and Ta’iz governorates, or indeed all of Yemen, and furthermore our estimates are subject to considerable imprecision and potential bias, highlighting challenges of primary data collection in insecure, politically contested settings (see below). The study does, however, indicate that community informants are able to recall details of local, adult decedents, even going back several years.

### Data collection challenges

Various challenges of this study arose during implementation. Limited electricity and internet coverage, ongoing SARS-CoV-2 transmission and unpredictable security conditions constrained data collection, especially in rural, remote sites within Ta’iz governorate, where the female researcher had to be accompanied by her spouse to secure travel. Moreover, some informants reported concerns that nominal details of decedents would be shared with warring parties, resulting either in non-participation or in short interviews. While informants generally recalled chronologically remote deaths, recall of ages and years of death was difficult, and informants appeared to mainly remember deaths during salient events (e.g. bombardments or battles) or decedents who are considered ‘martyrs’ in their community. This may explain the high proportion of war injury deaths among those reported. Capturing deaths among women and, in particular, children was difficult across sites, probably reflecting the choice of key informants (mostly men in positions of authority). Female researchers appeared better able to obtain this information from male and female participants, suggesting future similar studies in Yemen and comparable cultural settings should systematically deploy mixed-gender teams to collect data in communities and conduct formative work to ensure a balanced gender distribution among key informants. Lastly, entry and management of data on the ODK platform proved straightforward, though entering long lists of names onto smart phones was time-consuming.

### Limitations

Capture-recapture analysis involves certain statistical assumptions. Firstly, record linkage should be accurate, resulting in error-free attribution of decedents to one or more lists. Misclassification could have occurred due to the complexity of names in Yemen and the high frequency of certain names, combined with other errors in recall of key linkage variables. The long recall period affected recall of the exact full names of decedents by the key informants. If decedents are incorrectly classified as unique (i.e. less overlap among lists), overall mortality estimates would be upward-biased (over-estimated), while the inverse would happen if decedents on different lists are incorrectly matched. If misclassification is a function of date of death (e.g. more frequent errors for deaths in the remote past), our estimates of period mortality would tend to be artefactually higher in the pre-pandemic period than during COVID-19: this may be an explanation for the high pre-pandemic death rates in several sites, though it should be noted that data completeness was not correlated with date of death (data not shown).

A second assumption of capture-recapture methods is that lists do not draw on each other for information: this assumption is adjusted for in three- and four-list analyses, but may have biased our estimates for T1, T3, T4 and T5, where two-list analysis was done (in the latter, senior citizen and community leader lists may plausibly be non-independent).

Across all sites, we observed surprisingly little overlap among lists (Supplementary File), in contrast to previous applications of this method [30]. This may reflect individual informants being aware of deaths only within a sub-section of the site, difficulty in identifying all informants of the same typology within the site and/or local kinship, access to information, trust networks and ongoing insecurity.

Lastly, population denominators used to compute death rates are also subject to inaccuracy, not captured in the 95%CIs of our estimates. While all estimates were based on a 2004 census, projections rely on several statistical models and ground displacement information, all of which will typically have higher relative error at fine geographic resolution.

## Conclusions

This study provides some evidence of elevated mortality in parts of Yemen during the crisis period (particularly in communities with high death tolls due to war injuries), and of a mortality peak consistent with the first COVID-19 wave in 2020. However, study limitations considerably weaken this inference: as such, evidence from this study should be evaluated carefully alongside other efforts to document mortality and public health impacts of the war in Yemen. Future studies could more specifically compare sites affected directly by the conflict with those that weren’t.

This application of a key informant method to document mortality illustrates the feasibility and challenges of conducting such research in Yemen, and possibly comparable crisis-affected, insecure settings. We believe that key informant interviews combined with capture-recapture analysis could be applied in conflict-affected settings by taking into consideration the contextual differences and addressing the limitations we outline above. This method also has the potential to be applied prospectively as a substitute for non-functional vital events registration.

## Data Availability

Due to ongoing armed conflict and insecurity in Yemen, we believe that the benefit of publishing data, even if anonymised, is outweighed by the risk to participants. We may publish data in the future if the situation allows. All statistical code, and a dummy dataset that can be used to implement capture-recapture methods, is published at https://github.com/francescochecchi/mortality_capture_recapture_analysis

## Declarations

### Ethics approval and consent to participate

The study was approved by the Ethics Committee of the London School of Hygiene and Tropical Medicine (Ref: 22080) and Faculty of Medicine and Health Science, University of Aden, Yemen (REC-76-2020). Local approvals from the Ministry of Public Health and Health and Population Office in Ta’iz Governorate were obtained. All research participants provided verbal informed consents before the interviews.

### Consent for publication

Not applicable.

### Competing interests

The authors declare that they have no competing interests.

### Funding

The study was funded by the United Kingdom Foreign Commonwealth and Development Office, grant reference 300708-139. Funders were not involved in study design, data collection, analysis, or manuscript preparation.

### Authors’ contributions

Study design: FC, MA. In-country supervision: HB, FO, KZ. External supervision: HB. Data collection: MA, SA, HK, AB, YS. Data management: MA. Analysis: FC. Drafting manuscript: MA, FC. All authors commented on the draft and provided substantial inputs. All authors read and approved the final manuscript.

## Acknowledgments

We are grateful to all participants who gave their time to support this study, especially those in Yemen, who work and live under conditions of extreme hardship and adversity.

ODK servers & support were provided by the LSHTM Global Health Analytics Group (odk.lshtm.ac.uk).

## Notes

### Competing Interest Statement

The authors have declared no competing interest.

### Author Declarations

The study was approved by the Ethics Committee of the London School of Hygiene and Tropical Medicine (Ref: 22080) and Faculty of Medicine and Health Science, University of Aden, Yemen (REC-76-2020). Local approvals from the Ministry of Public Health and Health and Population Office in Taiz Governorate were obtained. All research participants provided verbal informed consents before the interviews.

