## Supplementary File for "Adult mortality before and during the COVID-19 pandemic in nine communities of Yemen: a key informant study"

#### Record linkage criteria

Table S1. Record linkage criteria and corresponding match classifications.

<table>
<tr><th colspan="3">Key variables</th><th colspan="4">Supporting variables</th><th rowspan="2">Overall match decision</th></tr>
<tr><th>Difference in names</th><th>Difference in year of death (y)</th><th>Difference in age at death (y)†</th><th>Gender</th><th>Address at death</th><th>Cause of death</th><th>Locality of origin</th></tr>
<tr><td rowspan="8">2-4 names available: all the same<br><br>OR<br><br>4 names: 3/4 are the same and the names are in the same order</td><td rowspan="4"><2</td><td><10</td><td>same</td><td>same</td><td>same</td><td>same</td><td>definite</td></tr>
<tr><td><10</td><td>same</td><td colspan="3">at least one variable is the same</td><td>probable</td></tr>
<tr><td>10-14</td><td>same</td><td>same</td><td>same</td><td>same</td><td>probable</td></tr>
<tr><td>15-19</td><td>same</td><td>same</td><td>same</td><td>same</td><td>possible</td></tr>
<tr><td rowspan="2">2-4</td><td><15</td><td>same</td><td>same</td><td>same</td><td>same</td><td>probable</td></tr>
<tr><td>15-19</td><td>same</td><td>same</td><td>same</td><td>same</td><td>possible</td></tr>
<tr><td>≥5</td><td><20</td><td>same</td><td>same</td><td>same</td><td>same</td><td>possible</td></tr>
<tr><td>any</td><td>any, but age ≥60yo at death</td><td colspan="4">not considered</td><td>possible</td></tr>
<tr><td>4 names: 3/4 are the same but the names are in different order<br><br>OR<br><br>3 names: 2/3 are the same and the names are in the same order</td><td><5</td><td><10</td><td colspan="4">not considered</td><td>probable</td></tr>
<tr><td>3 names: 2/3 are the same but the names are in different order</td><td><2</td><td><5</td><td colspan="4">not considered</td><td>possible</td></tr>
</table>

† If the age at death of either decedent being matched was <20yo, the cut-offs for tolerable differences were halved.

### Descriptive characteristics and list overlap by study site

#### Site A1

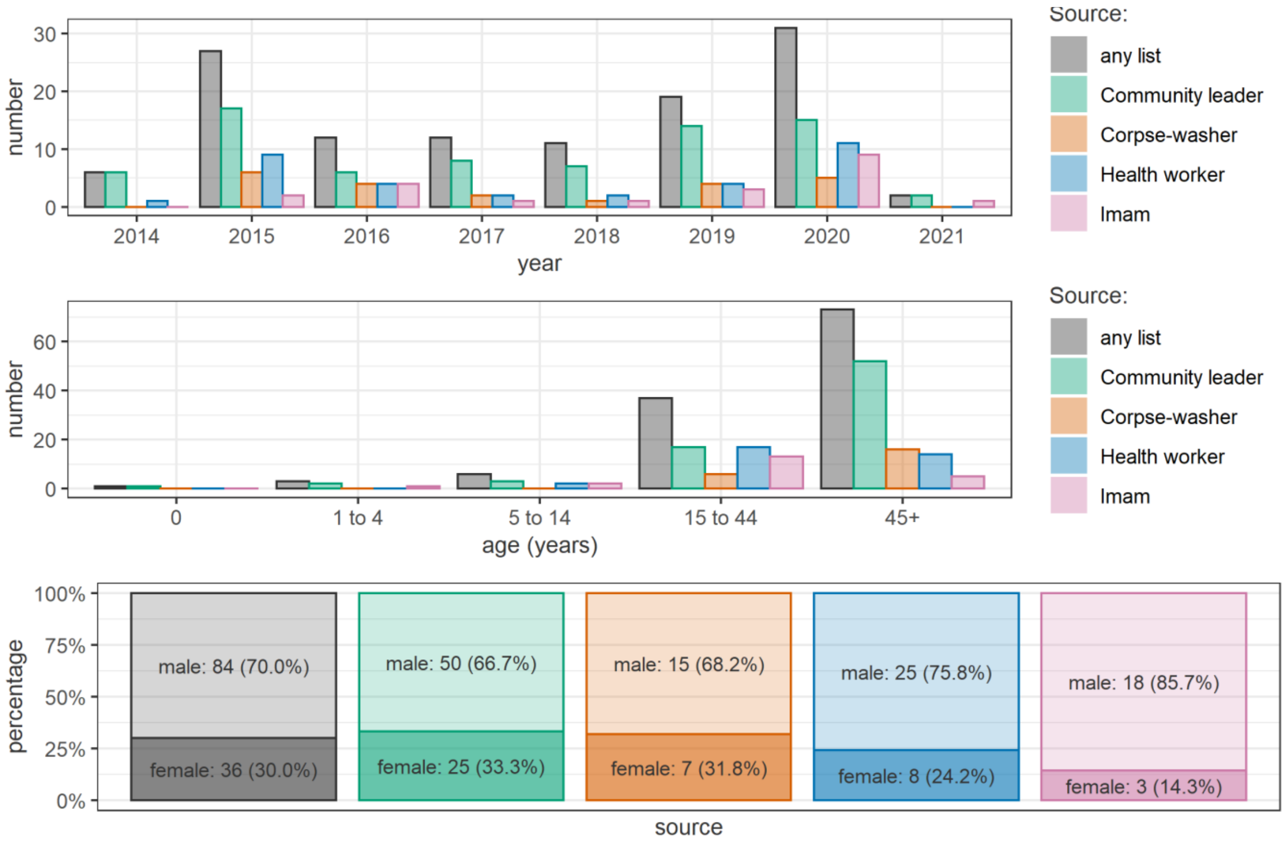


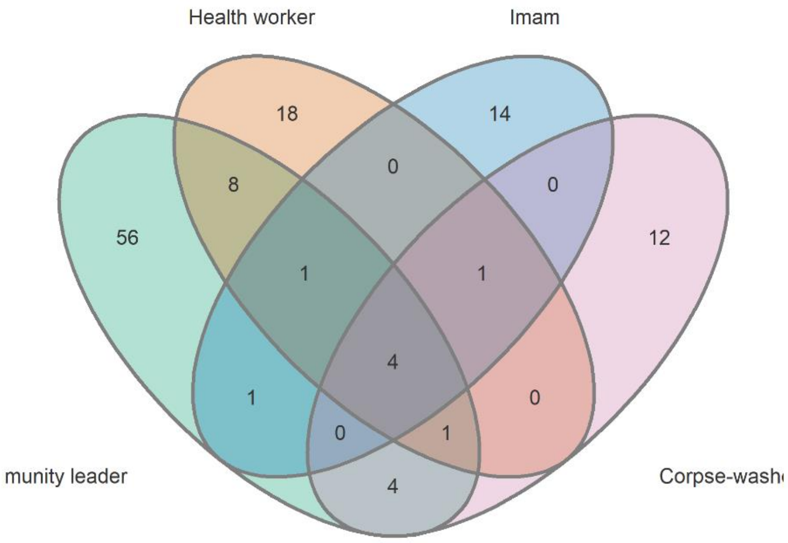

#### Site A2

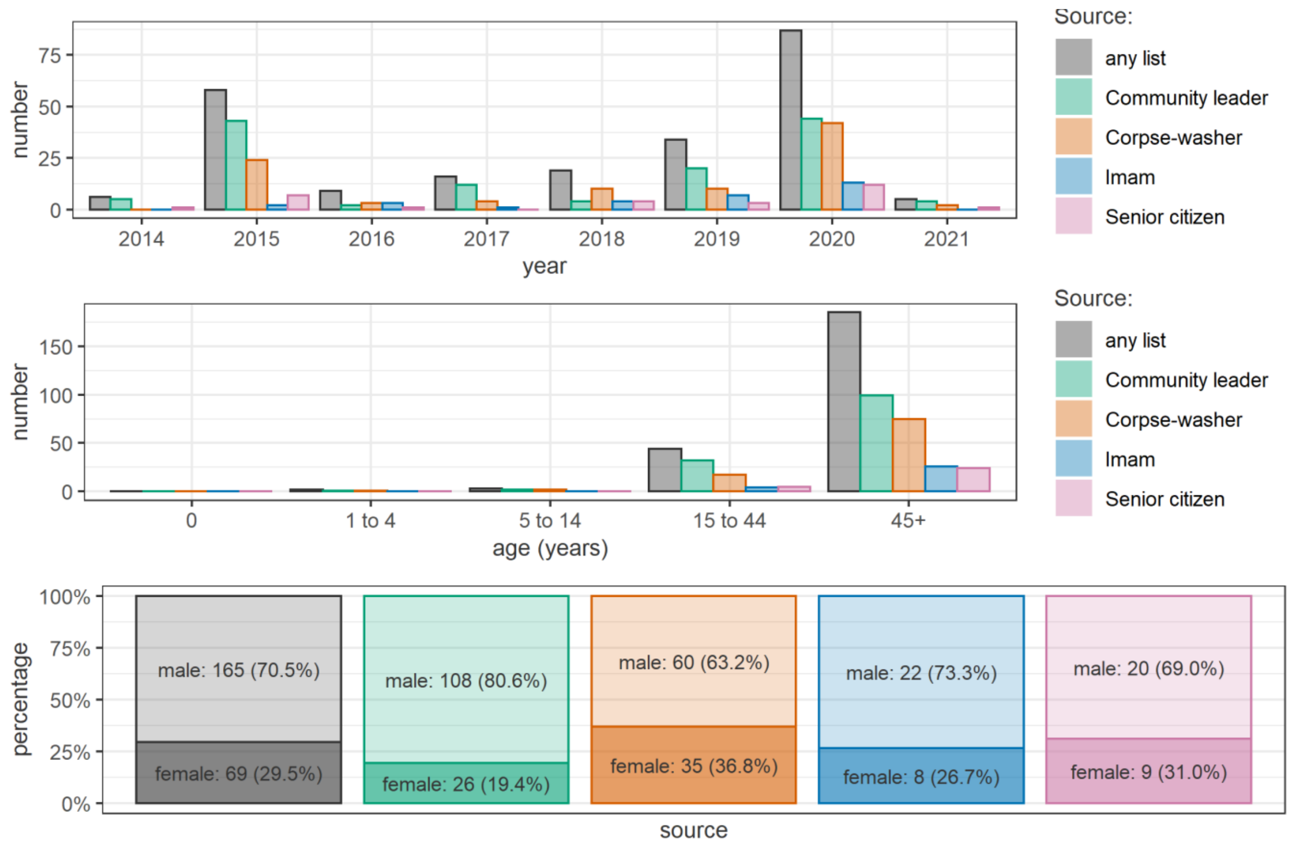
Source:
any list
Community leader
Corpse-washer
Imam
Senior citizen
number
year
2014
2015
2016
2017
2018
2019
2020
2021
age (years)
0
1 to 4
5 to 14
15 to 44
45+
percentage
source
male: 165 (70.5%)
female: 69 (29.5%)
male: 108 (80.6%)
female: 26 (19.4%)
male: 60 (63.2%)
female: 35 (36.8%)
male: 22 (73.3%)
female: 8 (26.7%)
male: 20 (69.0%)
female: 9 (31.0%)

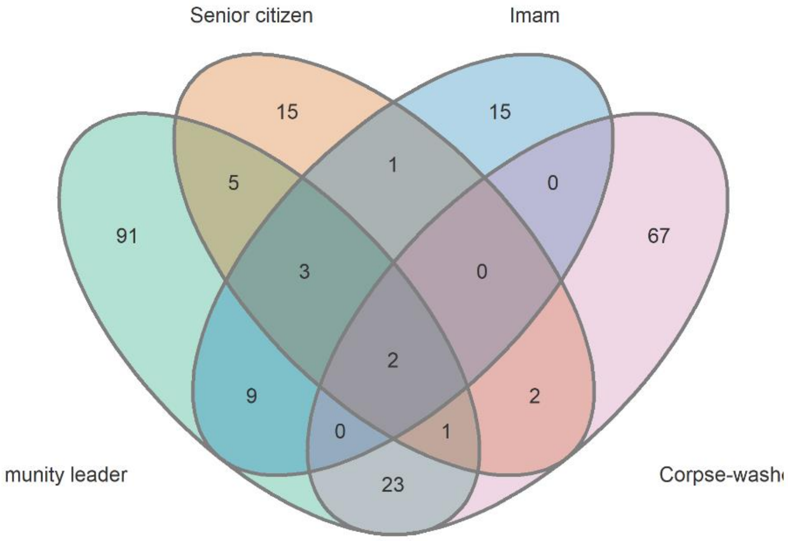
Senior citizen
Imam
munity leader
Corpse-wash
15
15
5
1
0
91
3
0
67
2
9
2
0
1
23

#### Site A3

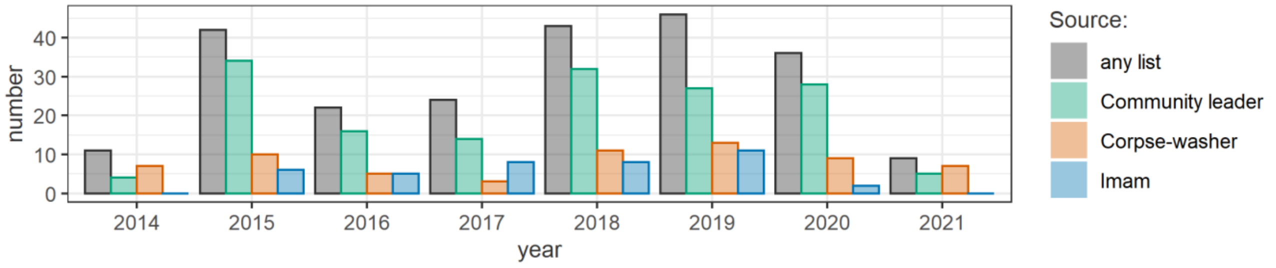
number
40
30
20
10
0
2014
2015
2016
2017
2018
2019
2020
2021
year
Source:
any list
Community leader
Corpse-washer
Imam

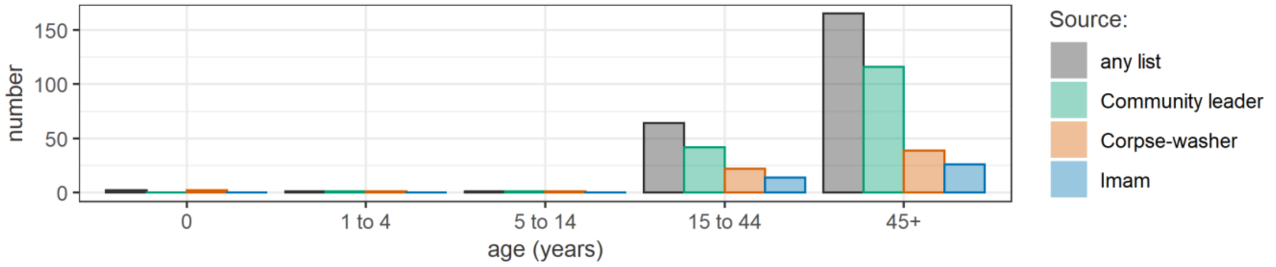
number
150
100
50
0
0
1 to 4
5 to 14
15 to 44
45+
age (years)
Source:
any list
Community leader
Corpse-washer
Imam

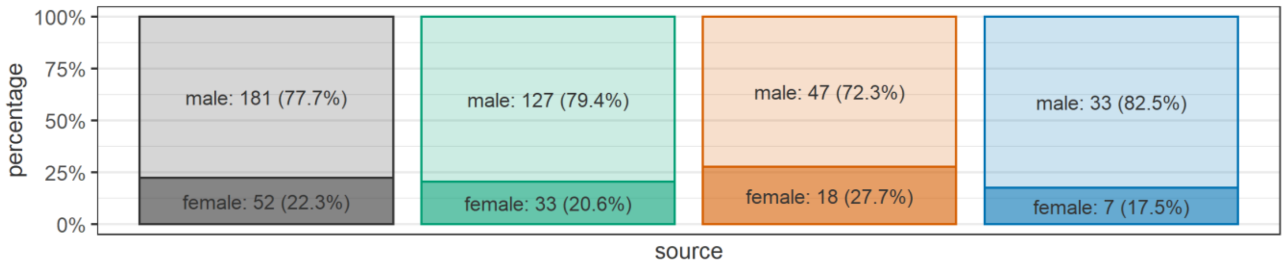
percentage
100%
75%
50%
25%
0%
male: 181 (77.7%)
female: 52 (22.3%)
male: 127 (79.4%)
female: 33 (20.6%)
male: 47 (72.3%)
female: 18 (27.7%)
male: 33 (82.5%)
female: 7 (17.5%)
source

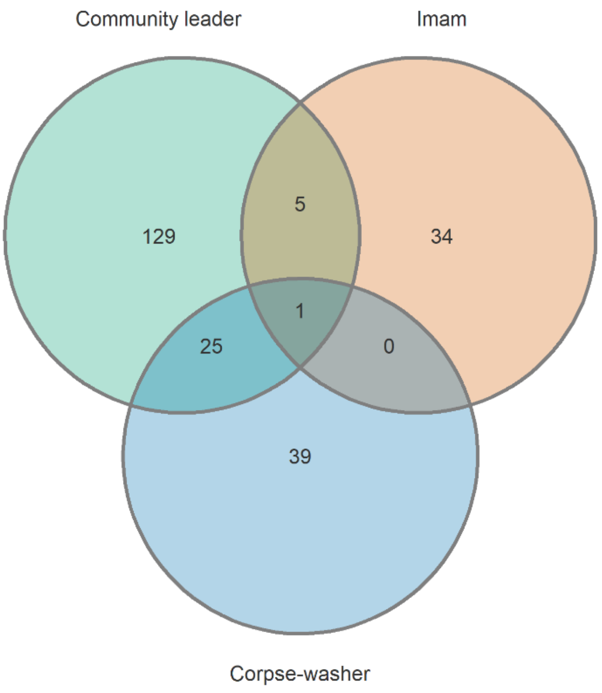
Community leader
Imam
129
5
34
1
25
0
39
Corpse-washer

#### Site A4

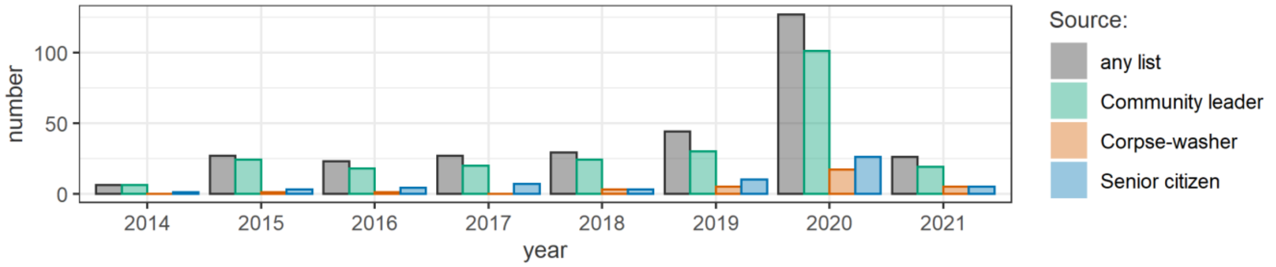

number
100
50
0
2014
2015
2016
2017
2018
2019
2020
2021
year
Source:
any list
Community leader
Corpse-washer
Senior citizen


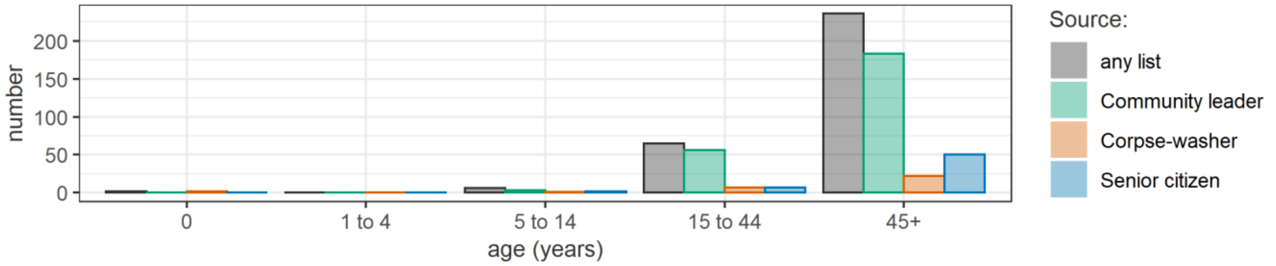

number
200
150
100
50
0
0
1 to 4
5 to 14
15 to 44
45+
age (years)
Source:
any list
Community leader
Corpse-washer
Senior citizen


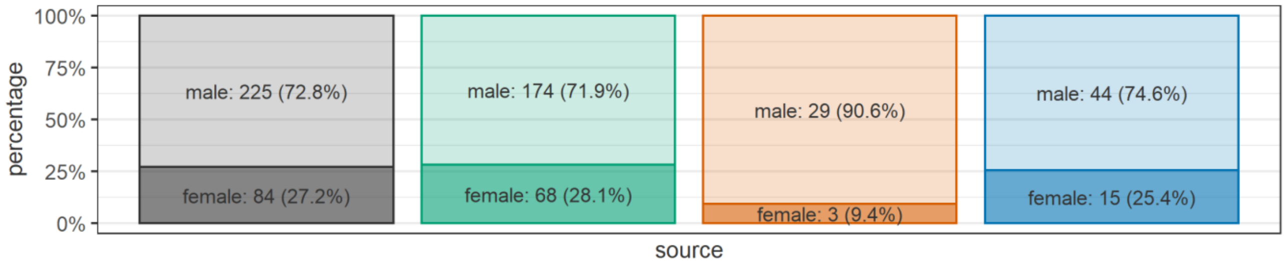

percentage
100%
75%
50%
25%
0%
male: 225 (72.8%)
female: 84 (27.2%)
male: 174 (71.9%)
female: 68 (28.1%)
male: 29 (90.6%)
female: 3 (9.4%)
male: 44 (74.6%)
female: 15 (25.4%)
source


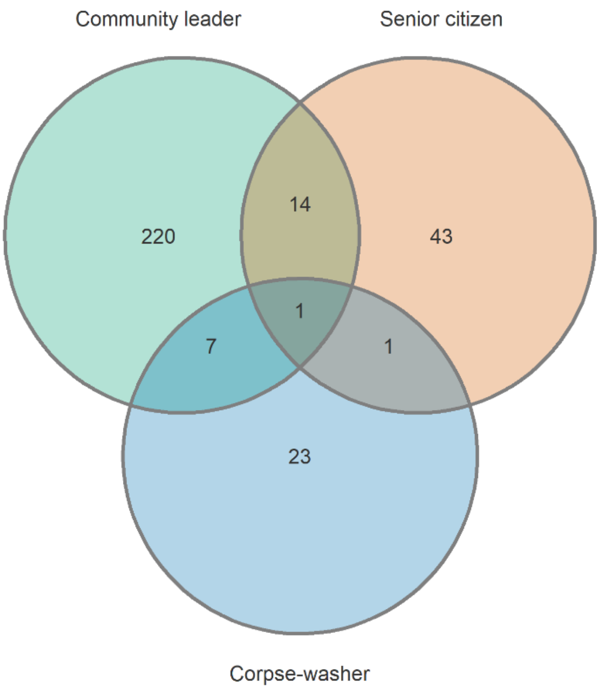

Community leader
Senior citizen
220
14
43
1
7
1
23
Corpse-washer

#### Site T1

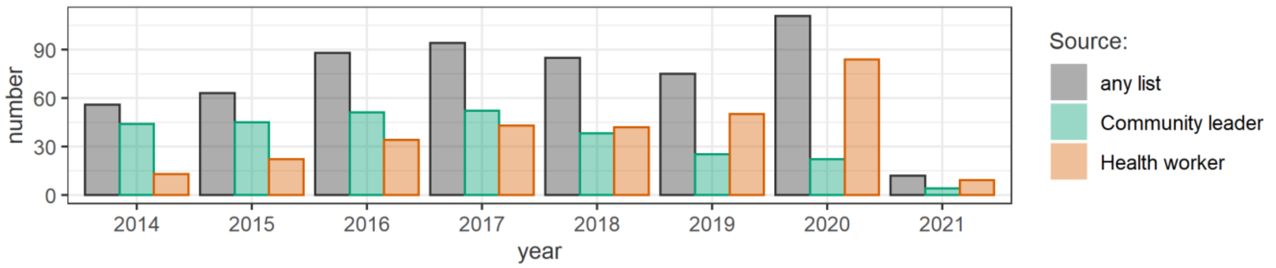
number
90
60
30
0
2014
2015
2016
2017
2018
2019
2020
2021
year
Source:
any list
Community leader
Health worker

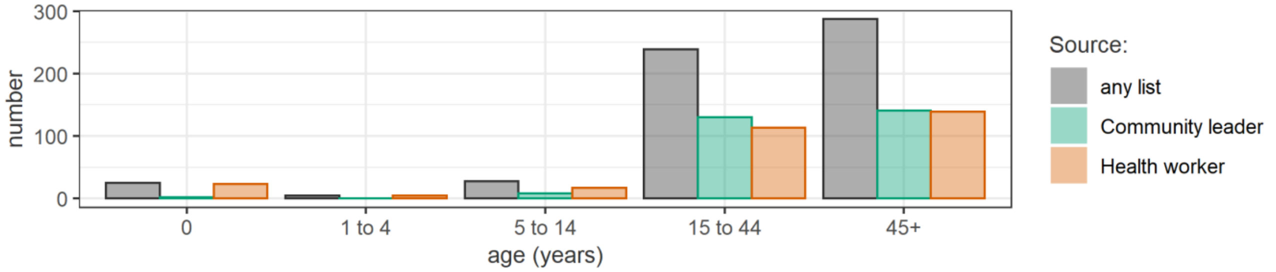
number
300
200
100
0
0
1 to 4
5 to 14
15 to 44
45+
age (years)
Source:
any list
Community leader
Health worker

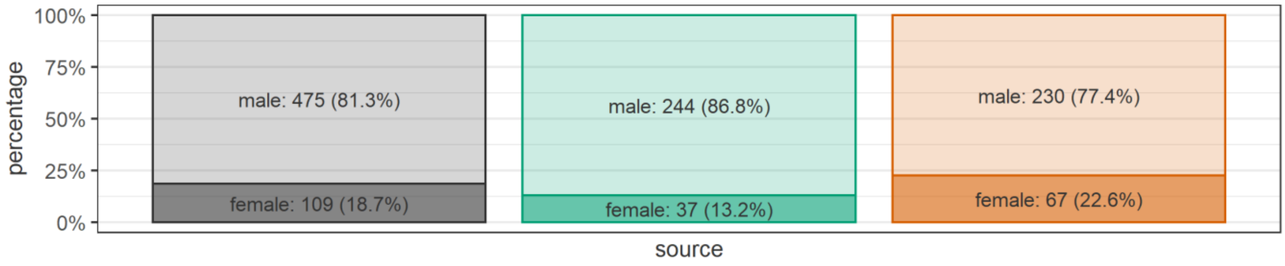
percentage
100%
75%
50%
25%
0%
male: 475 (81.3%)
female: 109 (18.7%)
male: 244 (86.8%)
female: 37 (13.2%)
male: 230 (77.4%)
female: 67 (22.6%)
source

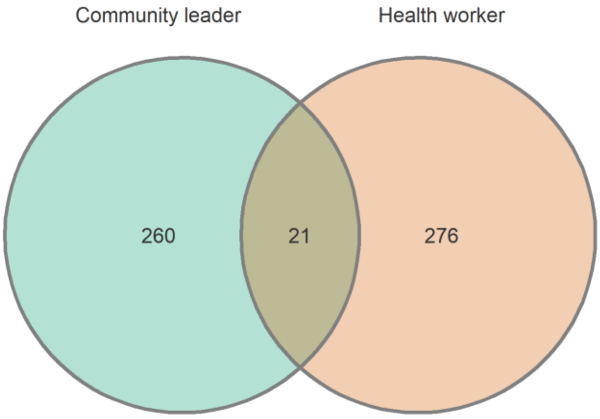
Community leader
Health worker
260
21
276

### Site T2

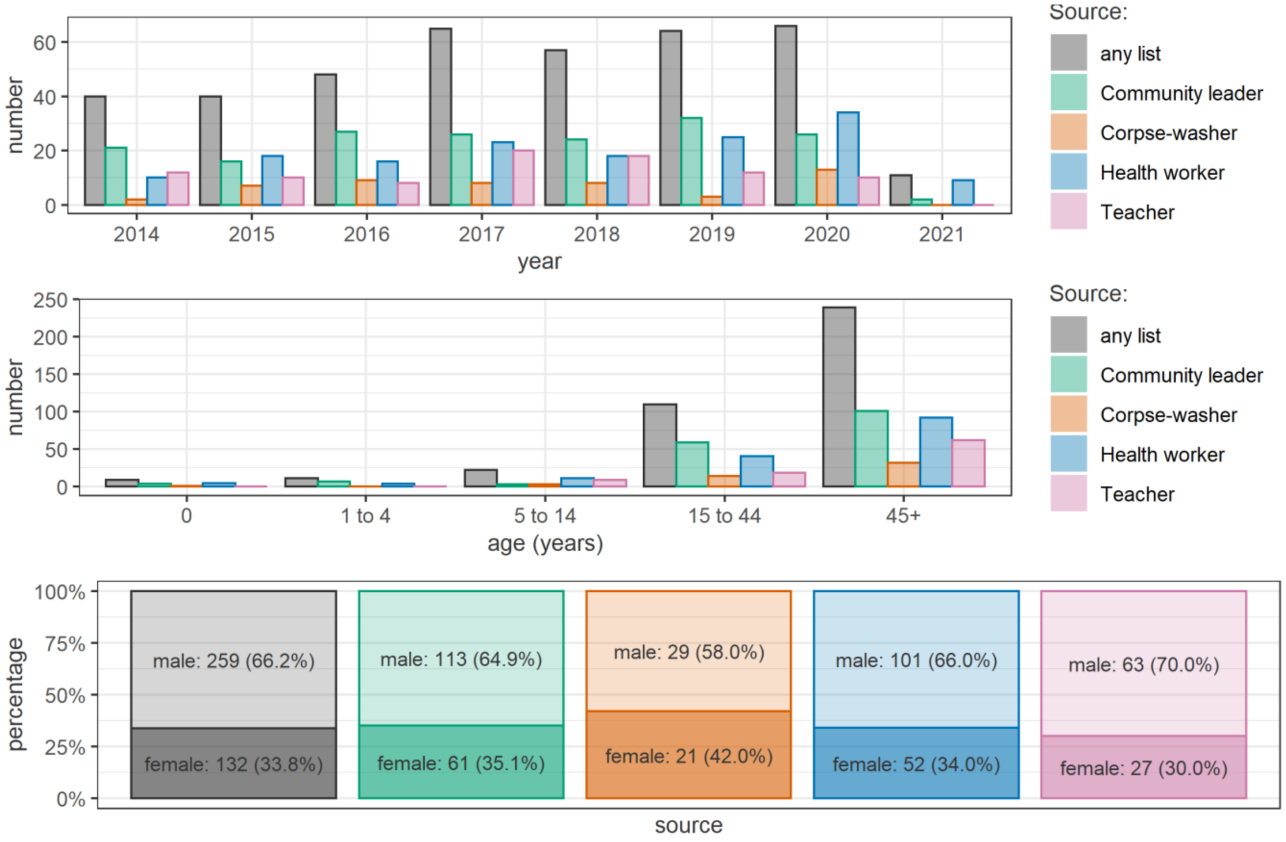
number
60
40
20
0
2014
2015
2016
2017
2018
2019
2020
2021
year
Source:
any list
Community leader
Corpse-washer
Health worker
Teacher
number
250
200
150
100
50
0
0
1 to 4
5 to 14
15 to 44
45+
age (years)
Source:
any list
Community leader
Corpse-washer
Health worker
Teacher
percentage
100%
75%
50%
25%
0%
male: 259 (66.2%)
female: 132 (33.8%)
male: 113 (64.9%)
female: 61 (35.1%)
male: 29 (58.0%)
female: 21 (42.0%)
male: 101 (66.0%)
female: 52 (34.0%)
male: 63 (70.0%)
female: 27 (30.0%)
source

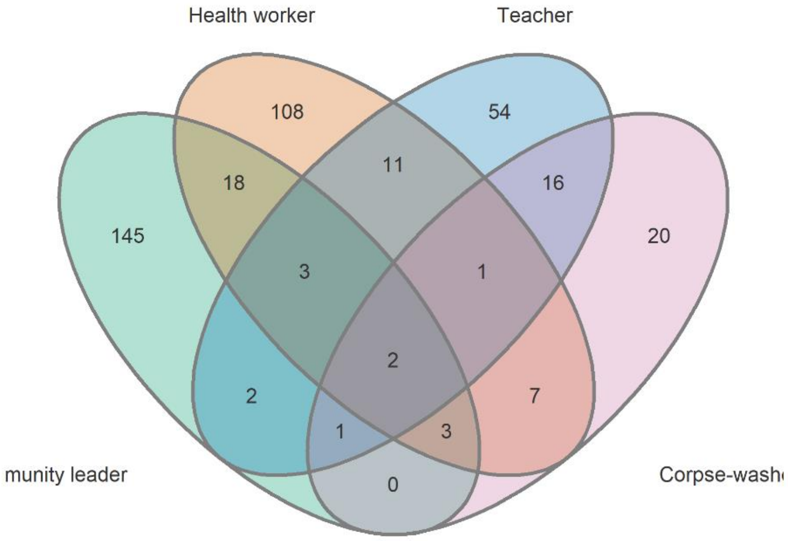
Health worker
Teacher
108
54
11
18
16
145
20
3
1
2
2
7
1
3
0
munity leader
Corpse-washe

#### Site T3

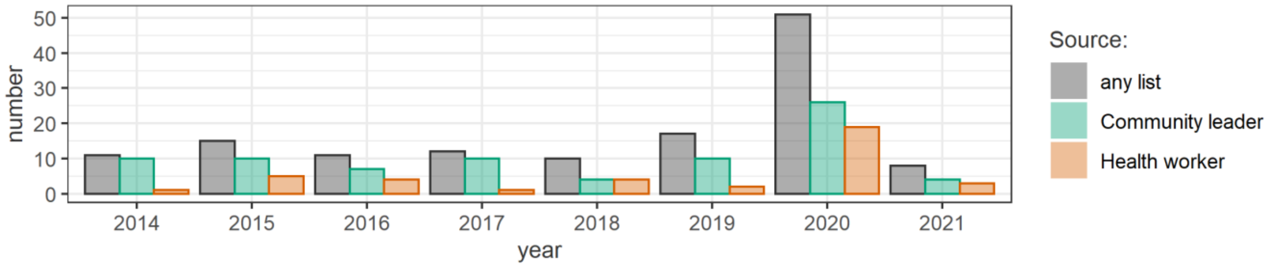
number
50
40
30
20
10
0
2014
2015
2016
2017
2018
2019
2020
2021
year
Source:
any list
Community leader
Health worker

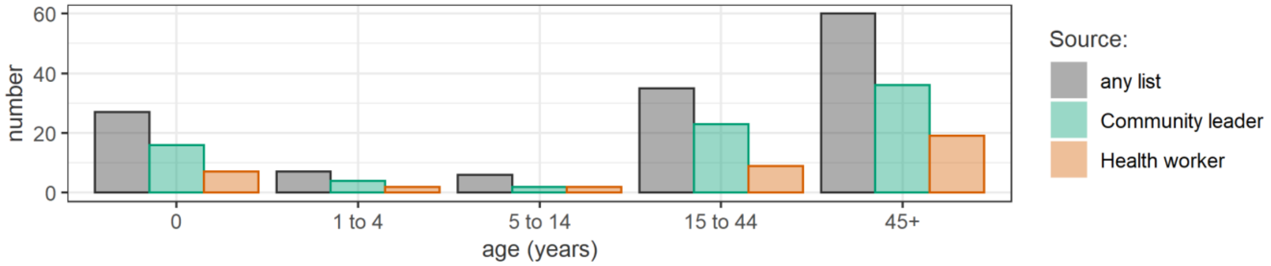
number
60
40
20
0
0
1 to 4
5 to 14
15 to 44
45+
age (years)
Source:
any list
Community leader
Health worker

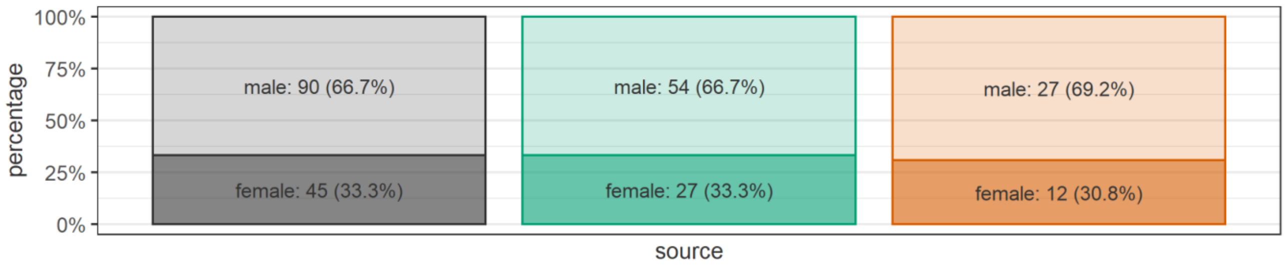
percentage
100%
75%
50%
25%
0%
male: 90 (66.7%)
female: 45 (33.3%)
male: 54 (66.7%)
female: 27 (33.3%)
male: 27 (69.2%)
female: 12 (30.8%)
source

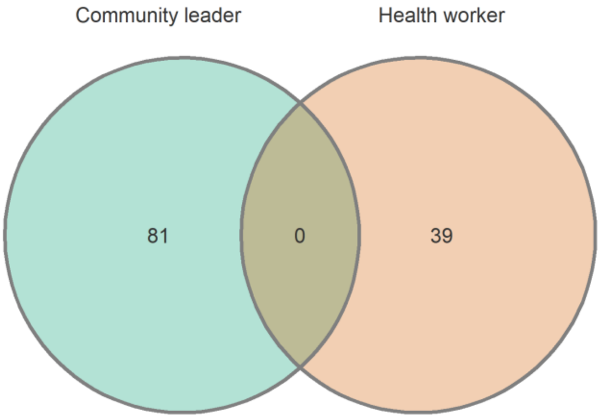
Community leader
Health worker
81
0
39

#### Site T4

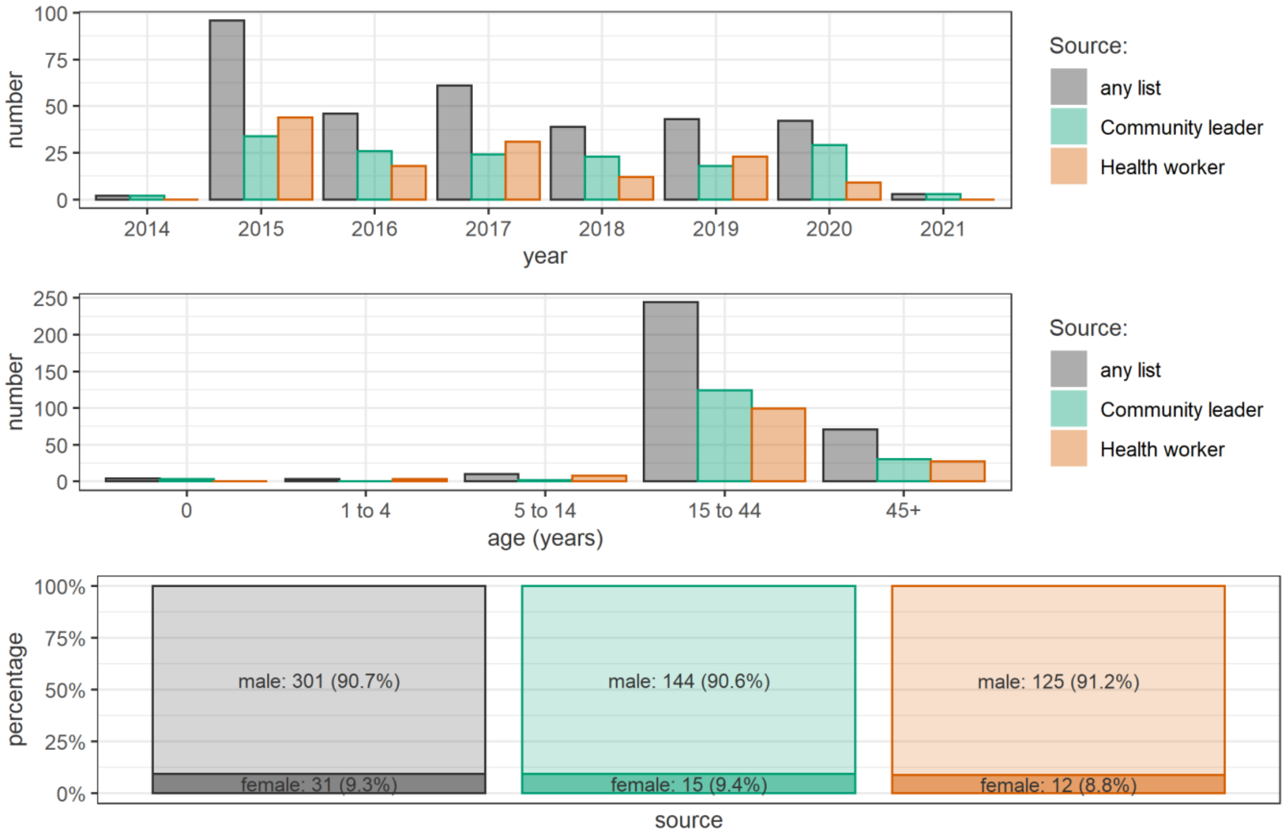

number
100
75
50
25
0
2014
2015
2016
2017
2018
2019
2020
2021
year
Source:
any list
Community leader
Health worker
number
250
200
150
100
50
0
0
1 to 4
5 to 14
15 to 44
45+
age (years)
Source:
any list
Community leader
Health worker
percentage
100%
75%
50%
25%
0%
male: 301 (90.7%)
female: 31 (9.3%)
male: 144 (90.6%)
female: 15 (9.4%)
male: 125 (91.2%)
female: 12 (8.8%)
source


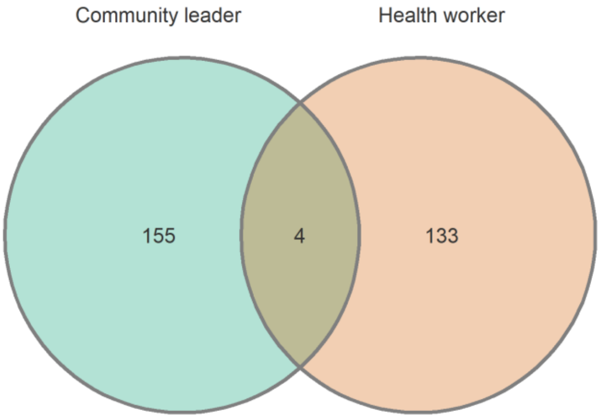

Community leader
Health worker
155
4
133

#### Site T5

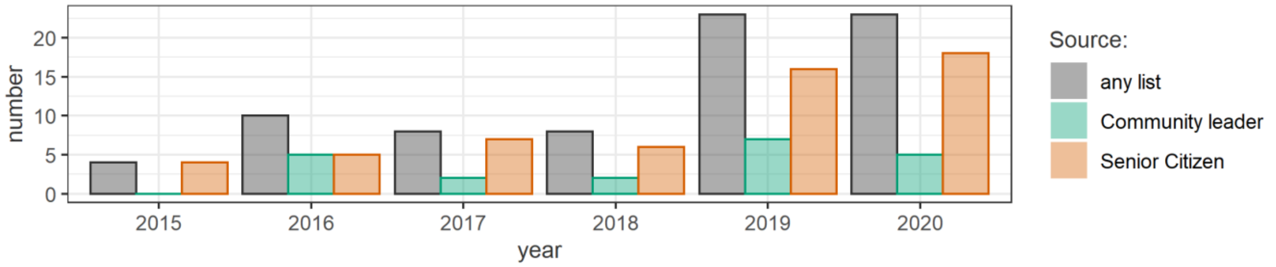
number
20
15
10
5
0
2015
2016
2017
2018
2019
2020
year
Source:
any list
Community leader
Senior Citizen

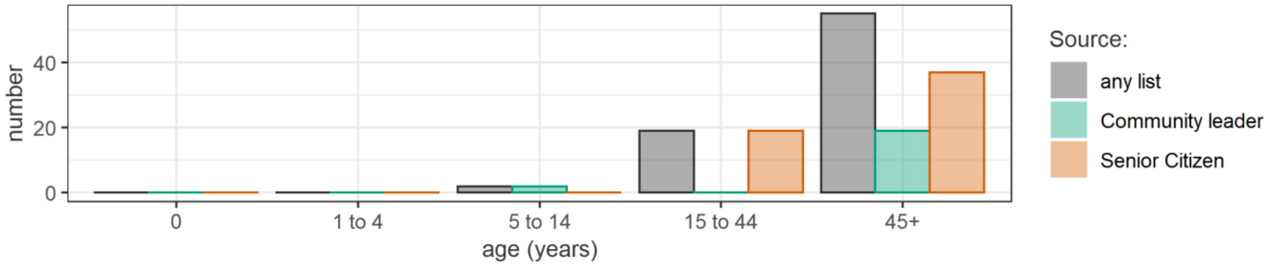
number
40
20
0
0
1 to 4
5 to 14
15 to 44
45+
age (years)
Source:
any list
Community leader
Senior Citizen

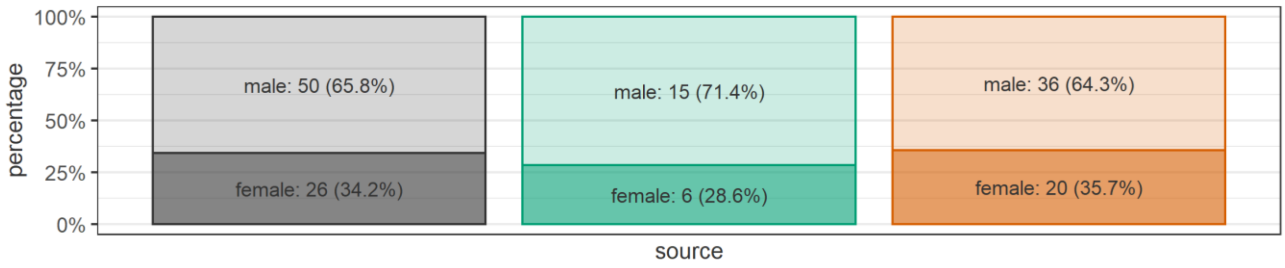
percentage
100%
75%
50%
25%
0%
male: 50 (65.8%)
female: 26 (34.2%)
male: 15 (71.4%)
female: 6 (28.6%)
male: 36 (64.3%)
female: 20 (35.7%)
source

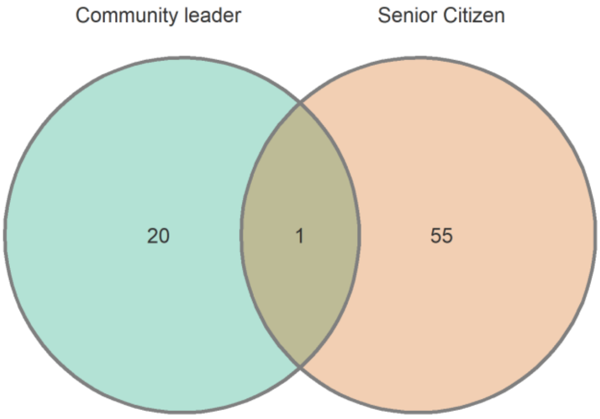
Community leader
Senior Citizen
20
1
55
